# Evidence-Based Complementary and Alternative Medicine Analysis of the current state of acupuncture clinical trial registration and reporting: for the past 9 years

**DOI:** 10.1101/2022.05.21.22275363

**Authors:** Quanai Zhang, Huijuan Lv, Yun Fu, Benlu Chen, Yingying Cai, Qiwen Zhang

**Author notes:** These authors contributed equally to this work (Quanai Zhang, Huijuan Lv).

## Abstract

**Background:** An increasing number of acupuncture clinical trials are registered, but the reports of results and data transparency are unclear. We aim to analyze and evaluate the current state of registration and reports of acupuncture clinical trials.

**Methods:** This paper focused on the acupuncture clinical trials that met the criteria in relevant studies registered and published during the period between 1 Jan 2013 to 31 Dec 2021. We traced by email to leaders that the report could not be searched. Besides, questionnaire investigation was chosen to acupuncture clinicians around by WeChat.

**Results:** The overall reporting rate of acupuncture clinical trial results from the two registration centers was 25%. Clinicaltrials.gov reporting rate was 12% and that from ChiCTR was 41%. Only a small proportion of trials were available on the registry website. Only the NIH clinical registry platform has records on the status of studies. We only received 3 replies that indicated the project was due to be ended but not completed. According to questionnaire, 157 clinicians have led clinical projects in Acupuncture. It shows that 84% projects were ended but not completed or not completed by the due date. About the reasons that caused the failure to find articles after the end of the project, 85% participants thought projects were ended but not completed or not completed by the due date. 15% participants thought projects were manuscript rejected, manuscript accepted but not in print, results protected, registration number unmarked, project delayed or negative results not submitted.

**Conclusion:** The number of acupuncture clinical trial registrations is steadily increasing, but the reporting rate of trial results is relatively low and the transparency of data is not ideal. Therefore, first of all, researchers should put more attention on trials designed, trial reported and respect to science. Secondly, set up a research progress tracker on registration platforms accessible to the public users. At the same time, the journal is suggested that adjust the evaluation criteria for reporting the negative results. Researchers with negative results are encouraged to show. Most importantly, the funders are recommended to optimize the ended review.

## Introduction

Clinical trial registration means that specific information about a trial needs to be prospectively registered in a web-based publicly accessible database [1]. Since the International Committee of Medical Journal Editors (ICMJE) announced in 2004 that clinical trials must be registered to be considered for publication [2], there has been an increasing emphasis on the regulation of acupuncture clinical trial registration to reduce the duplication of research [3] and to help prevent bias in reporting of results [4,5,6]. The Final Rule [7,8], which came into effect on 18 January 2017, provides further clarity on clinical trial registration and reporting requirements. The timely publication of trial data will facilitate academic communication, provide guidance on clinical practice and promote the development of evidence-based medicine [9].

After searching and collecting registered clinical trial literature, it is noticed that some of the clinical trial literature is marked with registration numbers, and some registered trials have not been published or data is unavailable. A study has evaluated the registration and reporting of clinical trials from the start of the registry to 2013 and the results were not optimistic [10]. To further explore the current state of the number of registered clinical trials on acupuncture and their reporting in recent years, this study intends to dig deeper into the clinical trials related to acupuncture registered from 2013 to 2021, in order to discover the problems and seek possible solutions to improve the quality of clinical research.

## Methods

### Search strategies

Terms, such as “needle” and “acupuncture”, are searched in Chinese Clinical Trial Registry platform[11]. We select “Advanced Search” with the search term “acupuncture” in Clinicaltrials.gov[12]. The search period was set from January 1, 2013 to December 31, 2021.

The publications were searched on the Web of Science, PubMed, CNKI, WanFang, VIP, and China Biomedical Literature Database, in English and Chinese only.

For some trials that the report could not be searched, we traced by email to leaders, the main issues investigated was the progress of trails.

Email response rate too low, so questionnaire investigation was chosen to acupuncture clinicians around by WeChat. The content of the survey was following: Question 1. Have you ever held a clinical project in Acupuncture? Question 2. Would your project be completed on time and would the results related to the research objectives be published before the end of the project? Question 3. What do you think caused the failure to find articles related to the research objectives in public available databases after the end of the project? (If the answer to question 1 is no, the rest of the questions cannot be seen).

#### Inclusion criteria

All clinical trials related to acupuncture (including electroacupuncture) registered between 1 January 2013 and 31 December 2021.

#### Exclusion criteria

Duplicate registered clinical trials.

#### Statistical analysis

After training three researchers responsible for the literature search (Benlu Chen, Yun Fu and Yingying Cai), data from the registered trials were extracted and summarized through EXCEL. And articles with published trial results were considered as results publication. The report was based on the above databases for subject terms, publication date, author, author institution, registration number, subject number, etc. If the publication is in the type of protocol or review, it is considered as other publication. The results of email responses and questionnaires are also summarized in Excel. For the extraction and searching process, if there is disagreement among three researchers, firstly, three parties should negotiate to solve the problem. If the agreement is not reached after the negotiation, then we need to seek help from experienced people or relevant experts.

## Results

The results of the search are shown in Fig 1. We found 2335 registered trials in ChiCTR and Clinicaltrials.gov. After selection, we finally selected 922 trials for our study. Of the included trials, ChiCTR had 166 trials whose results have now been published, and Clinicaltrials.gov with 62 trails. The results of the two centers are listed separately as there are differences in reporting and implementation of results and focuses on different issues. The state of trial registration and reporting is shown in Table 1.

**Fig 1.**
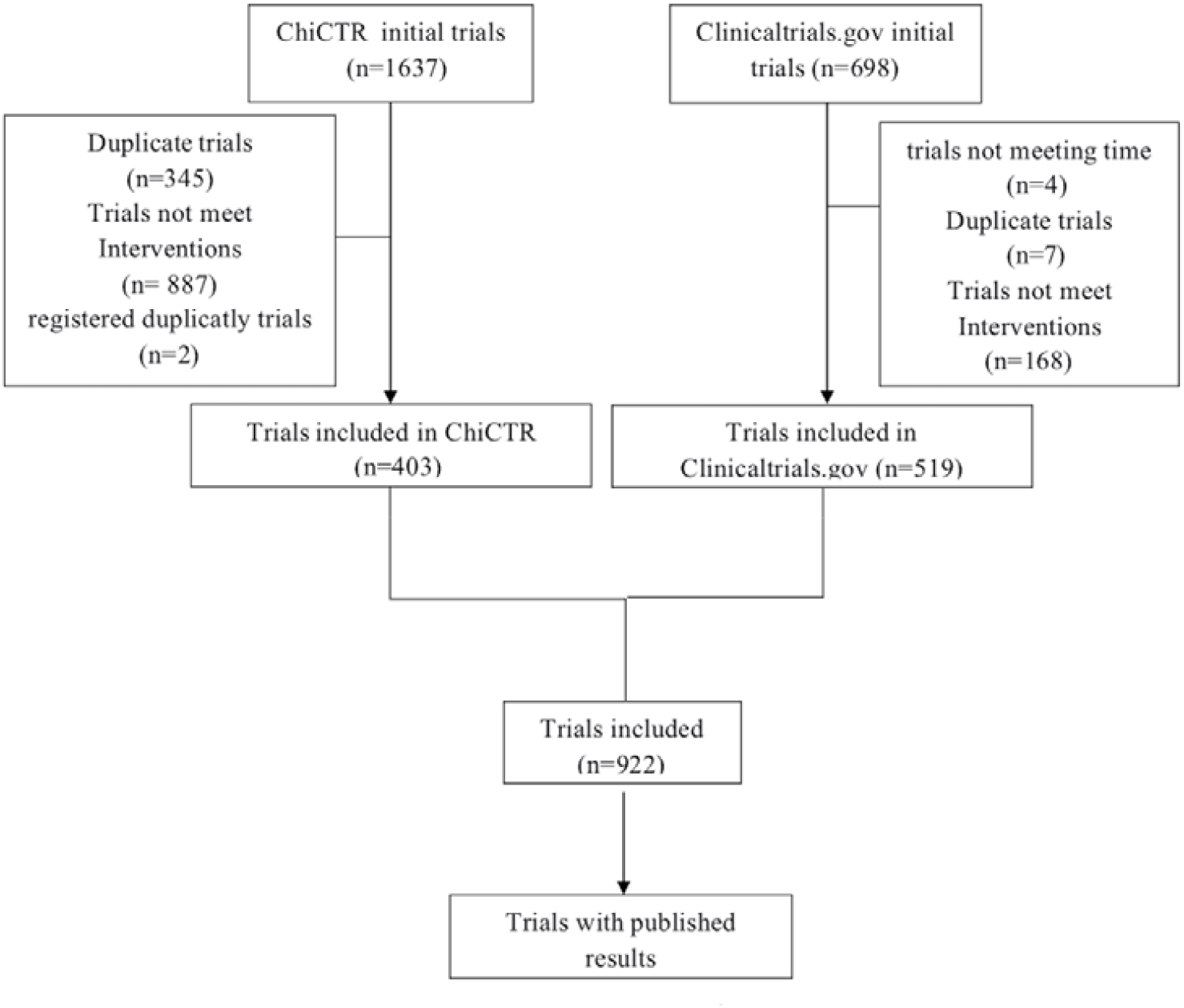
Flow chart of search

**Table 1.**
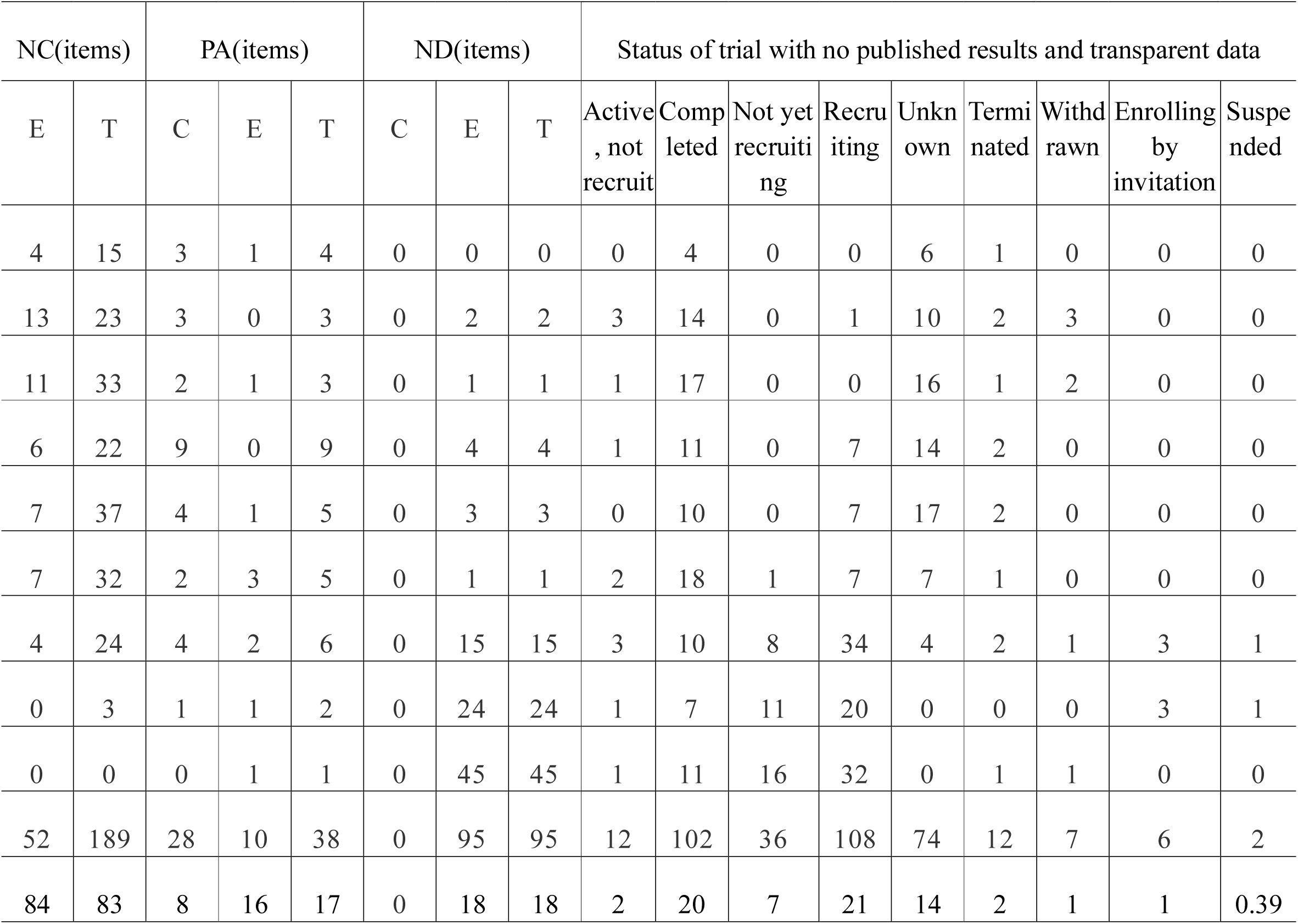
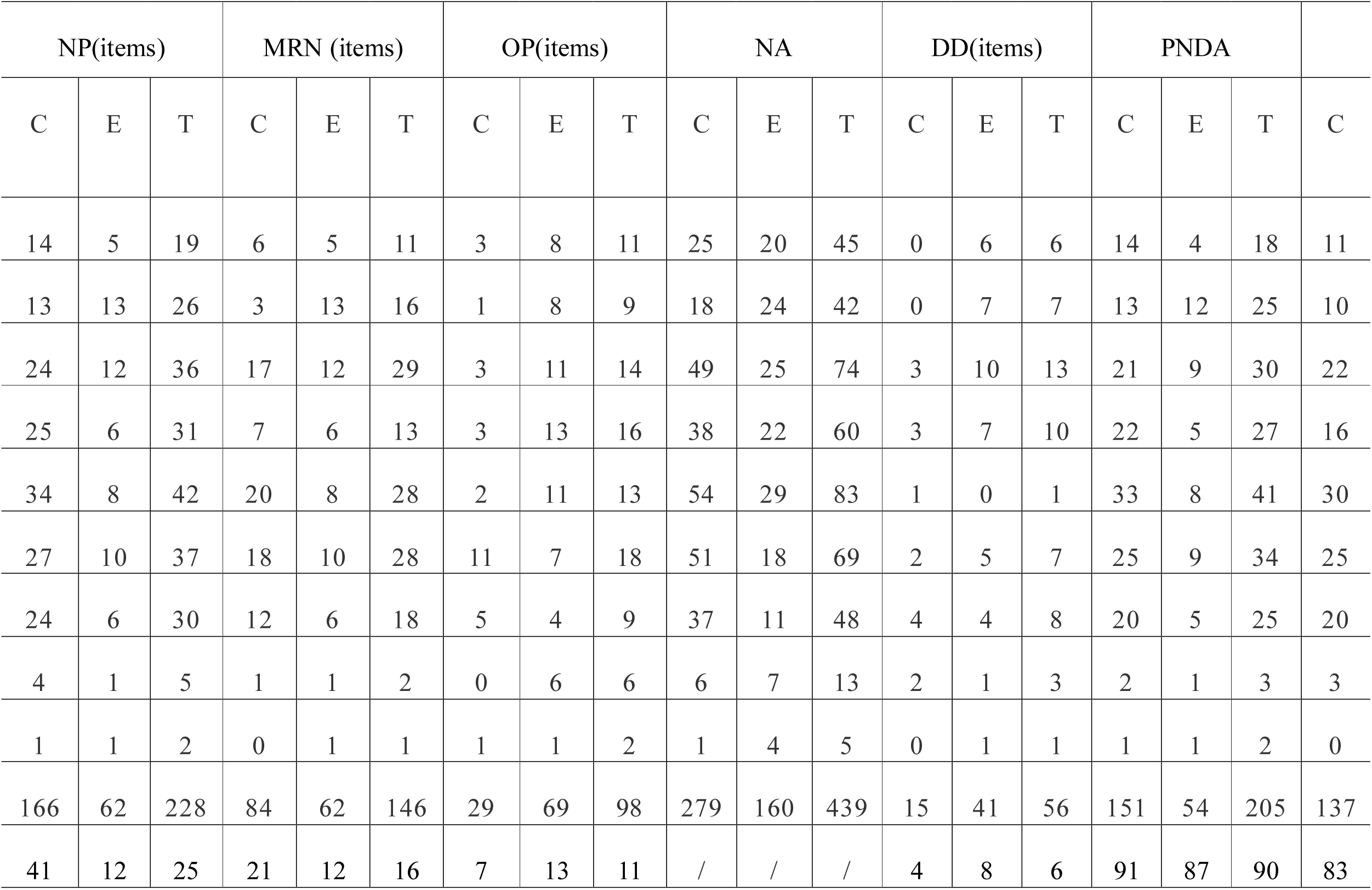
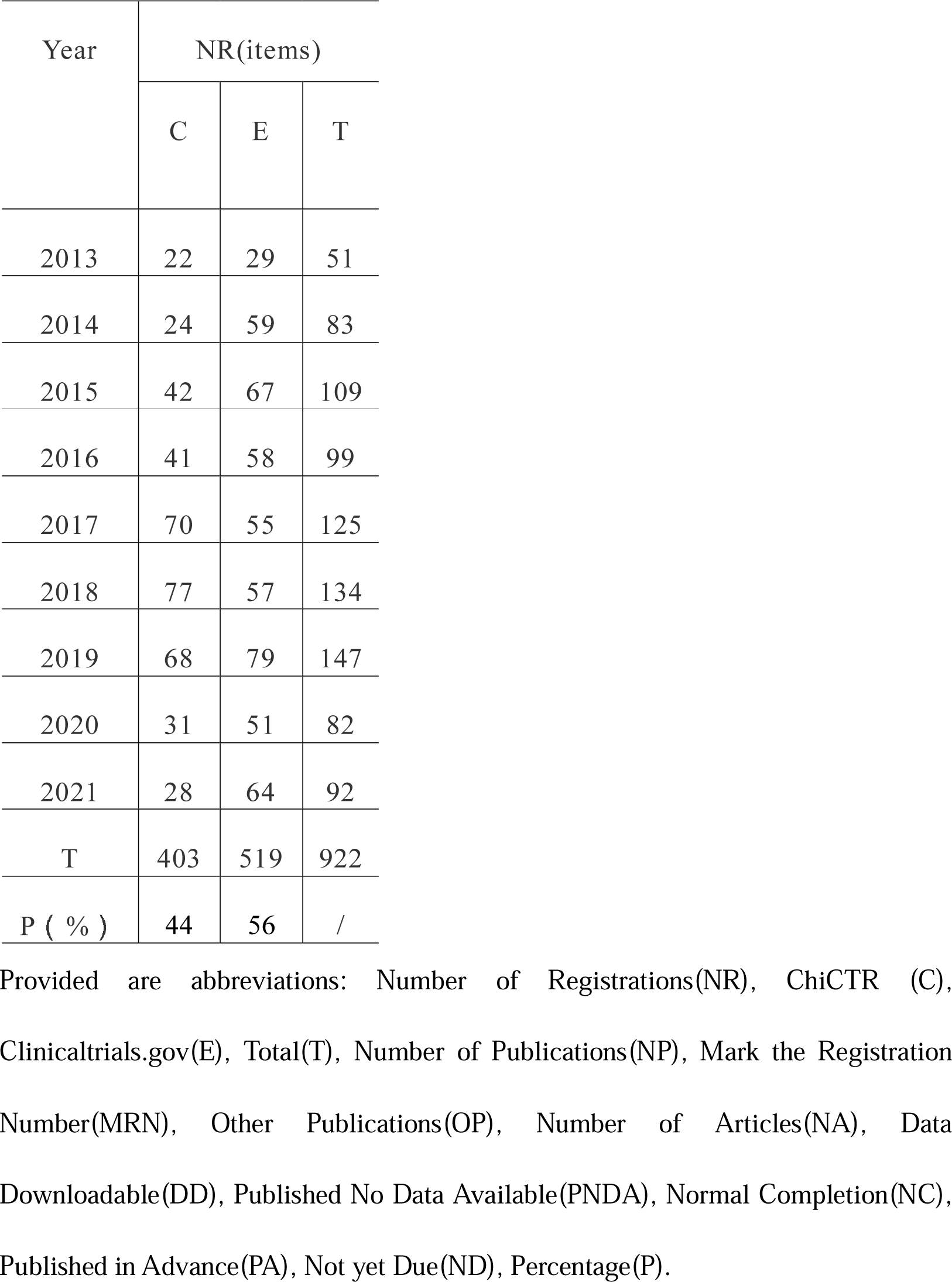
Trial registration and reporting state

### 1. Number of acupuncture clinical trial registrations

2014 witnessed the most significant increase in registrations of Clinicaltrials.gov. 2017 witnessed the most significant increase in registrations of ChiCTR and 2020 witnessed the felling.

### 2. Results publication rate

The total results reporting rate for acupuncture clinical trial articles in both registries was 25%. Specifically, Clinicaltrials.gov reporting rate was 12% for results, same as those reports with registration numbers marked. Other types of publication accounted for 13%. ChiCTR reporting rate was 41% for results, with 21% for reporting with registration numbers clearly marked, and 7% for other types of publication.

### 3. Publication timeline

The majority of trials with published results were published after completion of the trial, accounting for 83%. Specifically, Clinicaltrials.gov accounted for 84% of the trials registered; ChiCTR accounted for 83%.

Only 17% of the trials were published early. Of the unpublished trials, only 18% are in progress before the due date, while the rest were unpublished on due date.

### 4. Results data availability

Only a small proportion of trials were available on the registry website. A total of 6% trials were available from both registries. Specifically, 8% Clinicaltrials.gov trials were available; and 4% ChiCTR trials were available. 90% published trials’ data was not publicly available. Specifically, 87% published trials’ data of Clinicaltrials.gov was not publicly available and ChiCTR was available for 91%.

### 5. Study status of unpublished trials

Only the NIH clinical registry platform has records on the status of studies, of which the largest number were recruiting, completed and unknown details at 108, 102 and 74 respectively. A small number of trials were discontinued and suspended details at 12 and 2, respectively.

### 6. Survey results

We sent 694 emails but only received 3 replies. All email responses indicated that the project was due to be ended but not completed.

There were 218 acupuncture clinicians who took part in the questionnaire. 157 clinicians have led clinical projects in Acupuncture. But of them, only 12% projects were completed. 51% projects were ended but not completed. 33% projects were not completed by the due date. 4% projects were in progress. About the reasons that caused the failure to find articles related to the research objectives in public available databases after the end of the project, 52% participants thought projects were ended but not completed. 33% participants thought projects were not completed by the due date. 3% participants thought manuscript was rejected. 2% participants thought manuscript was accepted but not in print. 3% participants thought results need to be protected. 2% participants thought registration number was not marked. 3% participants thought project was delayed. 2% participants thought negative results were not submitted.

## Discussion

### 1. Registration trends

The number of acupuncture clinical trial registrations in both registries has steadily increased year by year. At the beginning of 2017, the fact that the number of ChiCTR registrations are significantly higher than the previous number, and even higher than Clinicaltrials.gov, is probably related to the application to WHO ICTR made by Clinicaltrials.gov in the US in 2017 and which is approved-local applications for registration when there is a WHO ICTR-accredited Level 1 registry where the trial is conducted [13]. There is a significant drop in ChiCTR registrations in 2020, likely due to the impact of the COVID-19 and the large amount of medical resources invested in China, but this does not affect the future trend of steady growth in acupuncture clinical trial registrations. Currently, acupuncture trials are in a period of modest growth, with no peak growth performance. Therefore, emphasis should be placed on clinical innovation and talent development in acupuncture, and researchers need to broaden the disease spectrum of acupuncture to better promote the development of acupuncture.

### 2. Reporting situation

Concerning the reporting of trials, it is clear that the reporting rates of results from acupuncture trials registered at both centers are low, and ChiCTR has a significantly higher rate of reporting results than Clinicaltrials.gov does. This may be related to the Chengdu Declaration published in April 2006, which announced the implementation of priority publication by ChiCTR member journals on a case-by-case basis [14]. There is no pattern in the number of trials published in terms of year due to a small number of publications. Based on the survey results, the low reporting rate is due to the following possibilities: some of trials were ended but not completed or not completed by the due date. Some of the trial manuscripts were rejected, which may due to low quality research or partial problems with the research program. Some of the trial manuscripts were accepted but not in print. Some of the trial results need to be protected. Some projects have been delayed. Some of the trials’ negative results were not submitted. In addition, it is possible that the low reporting rate is related to outcome reporting bias [15,16,17], and studies demonstrated that publication bias is a problem that still needs to be addressed [18].During the search process, some articles were found to be similar to the registered content but without the registration number to confirm. These articles rely on the responses of researchers to be sure, but most researchers did not respond to emails.

### 3. Data availability

The result of data availability in both centers is not satisfactory. According to our knowledge, some projects require that the results of the research be protected for a certain period of time. The data availability rate at Clinicaltrials.gov is significantly higher than that at ChiCTR, indicating that Clinicaltrials.gov is more concerned with the implementation of data disclosure. However, there is still much room for improvement in this area at both centers, far from the trial reporting rate. The centers need to play their roles as supervisors and ask trials that have published results to upload their data and take appropriate action if necessary. For trials with results that need to be protected, there is an option to first mark them on the registration website and make them public when the time comes.

### 4. Study Status

Clinicaltrials.gov clearly identifies trial study status and requires investigators to record any changes about trial status in the relevant section of the website. The main status can be divided by a few groups are Completed, Not yet recruiting, Recruiting, Enrolling by invitation, Active, not recruiting, Suspended, Terminated, Withdrawn and Unknown. The trials with the status of Terminated and Withdrawn could be conducted to improve the limitations of study field. In addition, it was found that most of the trials with the status of Completed had not been queried for publication. Maybe, some of which had been accepted but not in print and some of which were in the process of being written and some results were asked to be protected. Therefore, the status should be marked clearly, and measures need to be taken by the registry to urge and facilitate trials with the status of Completed to report their results and make their data publicly available. This shows that the research status of trials is of critical importance.

### 5. Proposed solutions

We need to face the problem of low reporting rates of registered trial results. Firstly, in response to the lack of registration numbers, trial centers and major journals should call for clinical trials to be registered before publication and require registration numbers to be indicated. Secondly, researchers are expected to pay more attention to the results reporting and data disclosure of registered acupuncture clinical trials. Thirdly, to reduce the likelihood of rejection, researchers need to be considerate when designing their studies and seek advice from experts. Then, registries will monitor the publication of results and timely updates in the registries. The registries shall, as far as possible, request investigators to update the progress of their trials, and shall take appropriate measures to promote the completion and reporting rate of trials that are not reported by the due date. At the same time, ChiCTR should be called upon to create a new research status item, and to set the number of years of research, and to urge investigators to update according to their research status. Funding institutions could also refer to the NIHR to request that registration information needs to be updated during the study period [19]. Furthermore, the potential for biased results reporting needs to be called out to journals and academics to pay more attention to the results of studies, rather than focusing on positive results. Researchers with negative results are encouraged to actively submit manuscripts. Last but not least, funding institutions should be strict in their project end approval, and could take a model from the NIHR [16] in requiring that the date of publication of trial results be disclosed and that past registrations and publications be taken into account when reviewing funding applications for new clinical trials.

### 6. Limitations

Limitations of this study: This paper mainly focuses on the acupuncture trials registered with Clinicaltrials.gov and ChiCTR, which may not reflect registrations and reporting from other registries, regions or countries. International studies may be carried out with global cooperation. Relevant information regarding WHO is not available which set a limit to data. The study is based on trials registered from 1 January 2013 to 31 December 2021. Not all leaders responded to the survey, which may cause bias in the results. The questionnaire was conceived with a small number of participants and should be followed up with a larger survey. Subject classification is not shown on the Clinicaltrials.gov website, hence, it is not possible to analyze the relationship between registration, reporting and subject classification.

## Conclusion

The number of acupuncture clinical trial registries is increasing steadily, but the reporting rate of trial results is very low and the transparency of data is not ideal. At the same time, the unavailability of registered trial results greatly weakens the advantages of clinical trial registries, causing a waste of research resources and funds and a possible shortage of research in the corresponding fields. Therefore, firstly, it is recommended that researchers need to put more focus on acupuncture clinical trial reports with respect and be considerate when designing their studies and seek advice from experts. Secondly, study progress can be recorded and monitored on the registration platforms which allow the public to access data. At the same time, the journal could update the evaluation criteria for evaluating negative-result reports to minimize the bias that may occur. Researchers with negative results are encouraged to actively submit manuscripts. Last but not least, funding institutions are expected to optimize the end-of-trial review, strengthen the reporting monitoring of core outcome indicators, regulate clinical research and reduce the waste of public resources.

## Data Availability

The data used to support the findings of this study are included in article. The original datasets presented in this study are available from the corresponding author on reasonable request.

## Data Availability

All data produced in the present study are available upon reasonable request to the authors

## Acknowledgments

This work was supported by the Zhejiang Provincial Administration of Traditional Chinese Medicine (grant no. 2020ZQ026) https://wsjkw.zj.gov.cn/, and the State Administration of Traditional Chinese Medicine of the People’s Republic of China (grant no. zyycx201901)http://www.satcm.gov.cn/.

## Author Contributions

Conceived and designed the experiments: QAZ and HJL conceived the ideas and drafted the manuscript. YF, YYC and BLC contributed to the acquisition and the analysis of data. QWZ revised the manuscript. All authors approved the final version of the manuscript accepted for publication.

## Notes

### Competing Interest Statement

The authors have declared no competing interest.

### Summary of Updates

Sections on methods, results, discussion updated to clarify conclusion.

